# Constructing cancer-specific patient similarity network with clinical significance

**DOI:** 10.1101/2023.05.05.23289558

**Authors:** Rukui Zhang, Zhaorui Liu, Chaoyu Zhu, Hui Cai, Kai Yin, Fan Zhong, Lei Liu

## Abstract

Clinical molecular genetic testing and molecular imaging dramatically increase the quantity of clinical data. Combined with the extensive application of electronic health records, medical data ecosystem is forming, which summons big-data-based medicine model. We tried to use big data analytics to search for similar patients in a cancer cohort and to promote personalized patient management. In order to overcome the weaknesses of most data processing algorithms that rely on expert labelling and annotation, we uniformly adopted one-hot encoding for all types of clinical data, calculating Euclidean distance to measure patient similarity, and subgrouping via unsupervised learning model. Overall survival was investigated to assess the clinical validity and clinical relevance of the model. Thereafter, we built a high-dimensional network cPSN (clinical patient similarity network). When performing overall survival analysis, we found Cluster_2 had the longest survival rates while Cluster_5 had the worst prognosis among all subgroups. Because patients in the same subgroup share some clinical characteristics, clinical feature analysis found that Cluster_2 harbored more lower distal GCs than upper proximal GCs, shedding light on the debates. Overall, we constructed a cancer-specific cPSN with excellent interpretability and clinical significance, which would recapitulate patient similarity in the real-world. The constructed cPSN model is scalable, generalizable, and performs well for various data types. The constructed cPSN could be used to accurately “locate” interested patients, classify the patient into a disease subtype, support medical decision making, and predict clinical outcomes.

## Introduction

Worldwide, there are approximately 19.3 million new cancer cases diagnosed every year[1]. Big data in cancer is being accumulated and reciprocally provides the opportunity to understand cancer more. While artificial intelligence (AI) is widely used in biomedical science[2], medical data analysis models based on other diseases or algorithms adaptive for other conditions are ineffective in the field of tumor, which hinders practical benefits of tumor clinical analytics. This is due to the granularity of clinical records is different in various disease[3]. Tumor histopathological data, molecular and genetic data represent pivotal features with high information density and clinical value[4, 5]. However, these two types of data are generally not involved in other diseases. Histopathological information is mainly clinical description, including imaging results, tumor site, tumor stage, differentiation, cellular composition, pathological type and final diagnosis. Molecular data comprises marker expression, genetic mutation, genomic features, and molecular classification. Through pathological examination and molecular detection, in combined with other clinical data, tumor and tumor microenvironment characteristics can be comprehensively described, and the exact diagnosis can be made, which underpin clinical decision making. Moreover, precision medicine claims making personalized therapy for every patient, fueled by clinical genetic testing because of advances in cancer molecular genetics and genomics [6].

Even now, cancer remains awkward and poses a great threat to human health. Inter-patient heterogeneity represents a great obstacle to cancer therapy. Conceivably, as cases of cancer are an enormous group, there are always some patients who are similar and historical similar patients may shed light on treatments for future patients. However, how to define and evaluate patient similarity remain controversial[7, 8]. Patient similarity calculation, which assesses the similarity between patients by mathematically calculating data on the multimodal heterogeneity metrics of patients, seems to be a solution. In general, the first step in patient similarity calculation is determining a multimodal data processing and integration strategy; The second step is to define a similarity metrics to calculate the distance or similarity score among patients in a systematic and consistent manner; The third step is to establish a patient similarity network (PSN), and carry out cluster analysis and clinical feature analysis in the PSN system; Finally, for a new patient to be evaluated, embed to PSN and define a group of patients most similar to the patient of interest in PSN based on the patient’s similarity score[9].

There were some explorations about patient similarity calculations in human diseases[7, 10]. They generally used patient demographic information, diagnosis, treatment, prescription drugs, laboratory test data, and physiological monitoring data extracted from electronic medical records (EMRs). At present some patient similarity calculations only use numerical variable for parameters to calculate Euclidean distance. This strategy presumes all variables are continuous, which is not perfectly suitable for categorical variables[11, 12]. Some use International Classification of Diseases (ICD) hierarchical coding to calculate the distance between the parent node and each child node for disease diagnoses, and then evaluate the similarity[13, 14]; Some orchestrate the medical record information into a medical knowledge graph, convert medical entity relation into vector space, which can calculate the Euclidean distance, Mahalanobis distance or Cosine similarity[15, 16]. The method of encoding/embedding conversion has obvious defects as that need to be converted into other systems such as ICD coding and knowledge graph, which are indirect calculation and bring various additional influencing factors and eventually affect the accuracy of the results.

Artificial Intelligence and machine learning have demonstrated usefulness in clinical data analysis[17, 18]. Diseases features are represented as vectors or matrices, and then artificial neural networks are used for similarity learning and patient clustering. The model obtained by deep learning algorithm is usually high specialized[19]. Most patient similarity models based on supervised or semi-supervised algorithms are dependent on pre-labelled training data and need extract parameters and corresponding exact weights. Although it performs well on the experimental dataset, the generalization ability is weak. Even for the same disease, the algorithmic model is difficult to transfer when their data metrics is different. This is drawback of supervised method[20]. Additionally, for defining and measuring patient similarity, data labelling is laborious and susceptible to subjective factors. In the era of biomedical big data[21], knowledge and decision are obtained based on population data but not on clinician’s experience, so the labelling is disgusted. Temporarily these two shortcomings lead to little clinical application value of deep learning represented by supervised neural networks in patient similarity assessment, self-organizing map is a promising neural network besides.

In view of the above considerations, we employed one-hot encoding and unsupervised clustering to manipulate heterogeneous clinical data to assess the similarity of tumor patients. The constructed PSN was validated by survival endpoints to make sure clinical validity. Our cPSN would create paths from clinical data to insight, from information to decision. With an emphasis on clinical utility and usability, clinical investigators can use cPSN to find insights and conduct clinical research. Clinicians can use cPSN to inform patient stratification, to recommend treatments, to deliver personalized patient care, and then to improve population health management[22, 23].

## Methods

### Data collection and preprocessing

One thousand patients of surgical gastric cancers (GCs) with multiple types of clinical data were collected from the department of gastrointestinal surgery, Shanghai Changhai Hospital. Clinical information was extracted from EMRs, medical examination reports, and then data was preprocessed to ensure consistency in formatting. Clinical descriptions were summarized into keywords, such as classifying surgical procedures into laparotomy or laparoscopy. For histopathological data, our dataset contained mesenteric vein/portal vein involvement, qualitative description of surgical margin status, tumor stage, tumor differentiation and so on (Table 1). For molecular genetic data, the dataset contained gene mutation derived from clinical genetic testing, gene expression and immunohistochemical data. Emphatically each gene mutation and each tumor marker/gene expression level can be considered as an independent variable. Missing data are padded with NA.

**Table 1.**
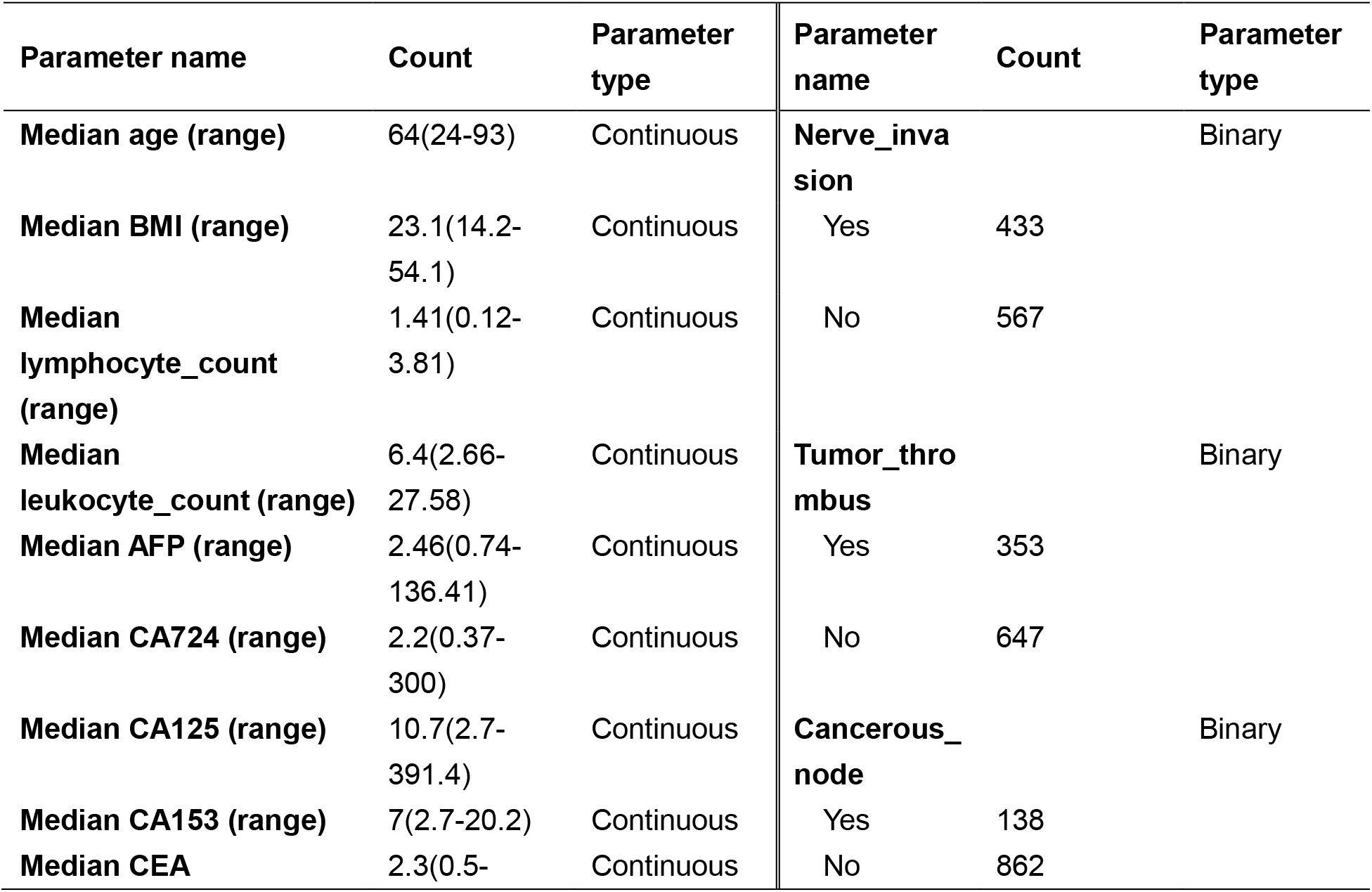
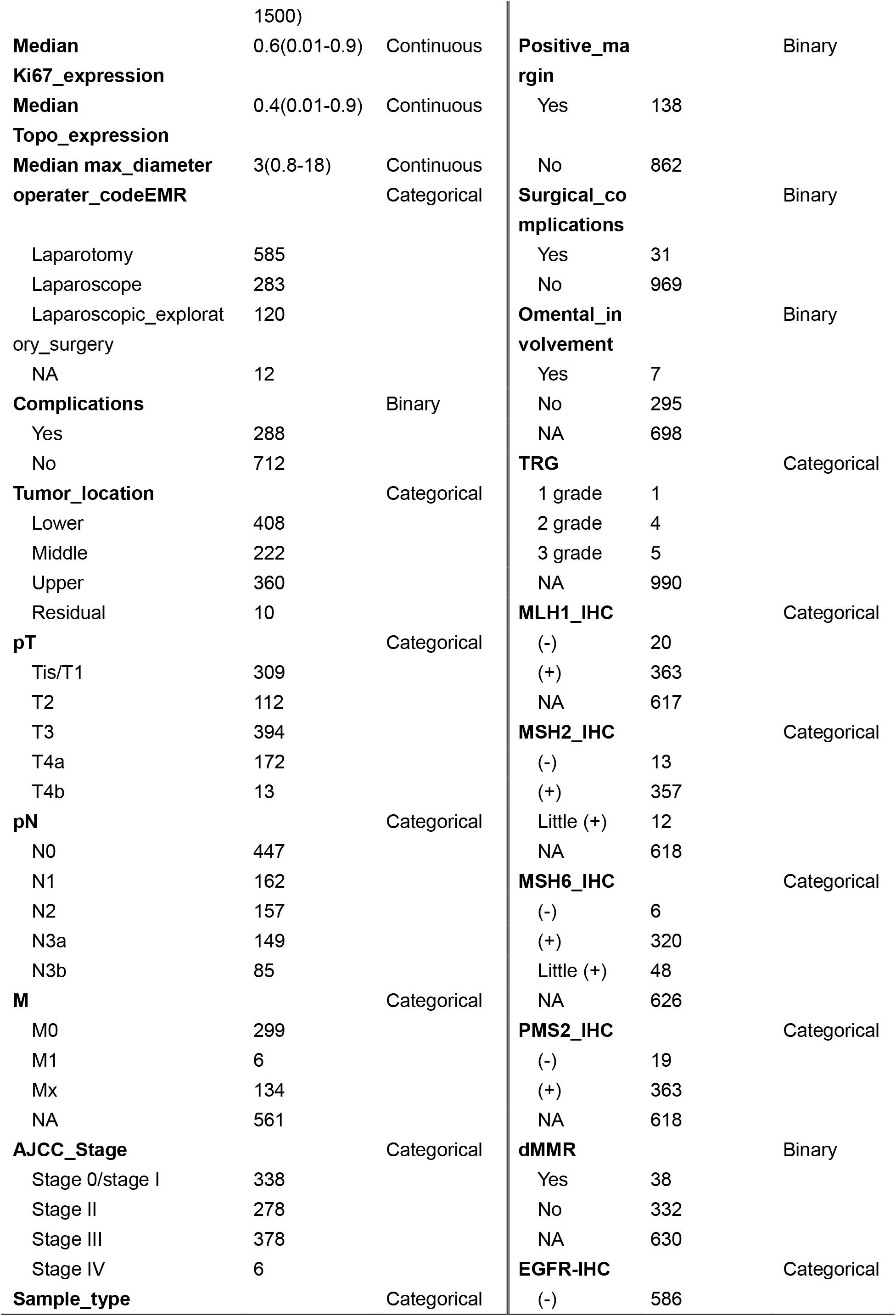
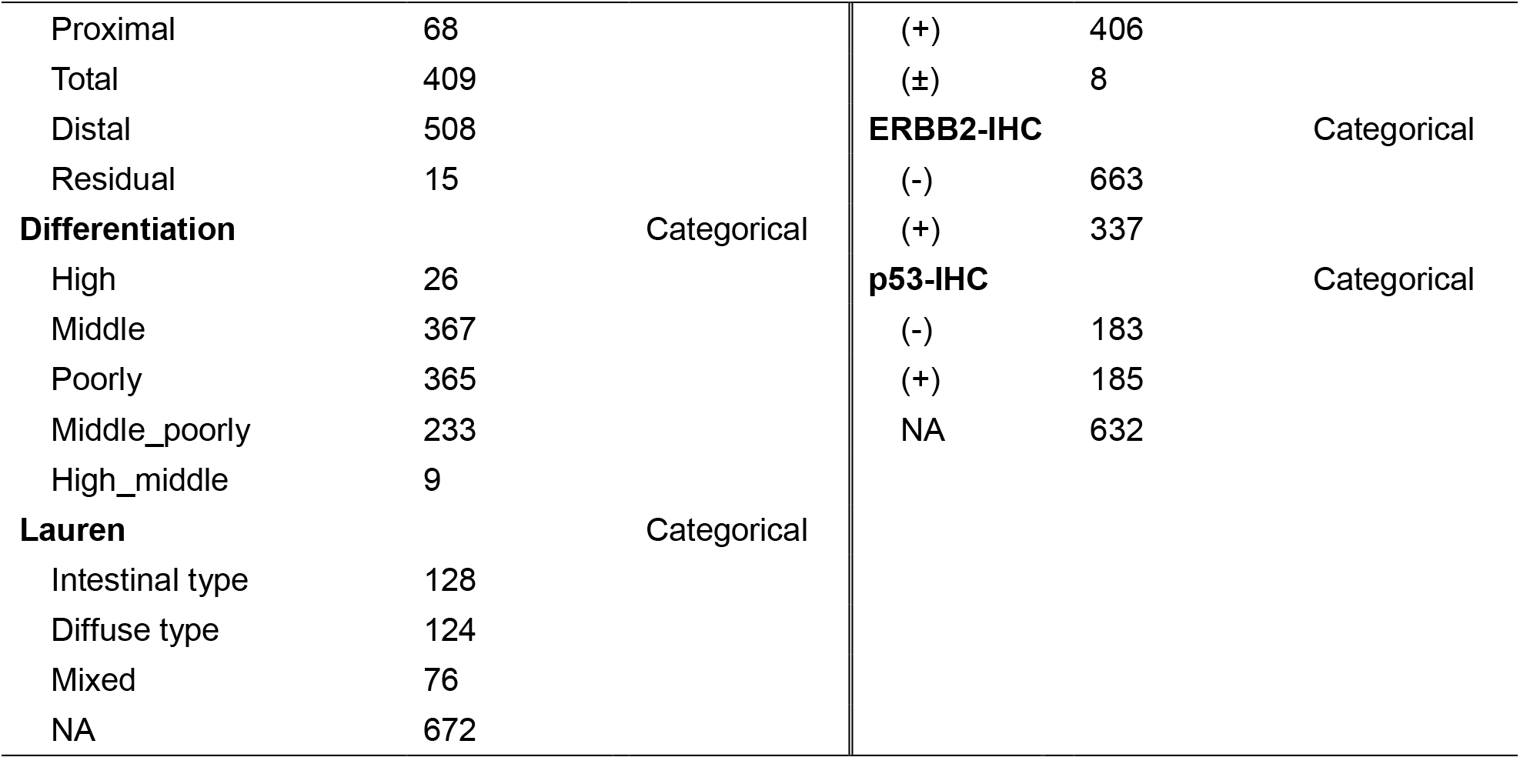
Data summary of baseline characteristics of GCs patients

### Encoding

In the encoding process, categorical variables were directly coded, numerical variables and clinical qualitative descriptions were converted into categorical variables, and each categorical state of each variable was recorded as a one-hot feature. Supposed that there are *M* observation indeces (variables) in a set of samples, denoted as,*X*_1_,*X*_2_,…,*X*_*M*_, and each observation index *X*_*i*_ has *N*_*i*_ different classification states, denoted as *N*_1_,*N*_2_,*N*_*M*_, altogether obtained 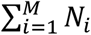 one-hot features. Continuous values were transformed into discrete values by equivalent partitioning. Preferably, for numerical variables, the values in a set of samples were divided into 4 parts according to the quartile method so that 4 categorical variables were formed. For clinical qualitative descriptions, *N* states formed *N* categorical variables.

The missing value was regarded as an independent one-hot coding type in the observation index of clinical data, and there is no need to fill in null values.

One-hot encoding method was engaged to integrate multimodal medical data. Subsequently, the heterogeneous data of patients were transformed into a feature embedding matrix.

Through distance calculation (Euclidean distance in this study), the feature embedding matrix was orchestrated to a PSN. Preferably, the *t*-distributed stochastic neighbor embedding (*t*-SNE) method can be used to visualize the high-dimensional network, in the way of two-dimensional or three-dimensional display.

### Subgrouping

*K*-Means clustering, an unsupervised learning algorithm, is conducted for patients’ similarity analysis to divide all patients into *K* clusters. *K* is a positive integer greater than or equal to 2. If *K*=2, there will be two nonoverlapping clusters. The elbow method or gap statistic method is used to evaluate the effect of clustering for each selected number of *K* clusters. Data encoding and clustering analyses were conducted using scikit-learn packages in python3.10.

### Survival analysis

The Kaplan-Meier method was used for clinical endpoint correlation analysis. The log-rank test was used to assess the statistical differences between overall survival (OS) of different groups of patients after clustering. PSNs with or without clinical implications were obtained based on the statistical significance of *p*-values. If the p-value less than 0.05 we would consider the constructed PSN is correlated with clinical meaningful endpoint, namely cPSN. Survival analysis was conducted in R4.0.3 using the survival and survminer packages.

## Results

We collected 1,000 surgical gastric cancers with multiple types of clinical data (Figure 1). In this study, the heterogeneous medical data we dealt with included demographic data, histopathological data, molecular and genetic data, laboratory tests and the surgical paradigm narrative. The types of data contained numerical variables, binary variables, categorical variables and clinical qualitative descriptions (Table 1).

**Figure 1.**
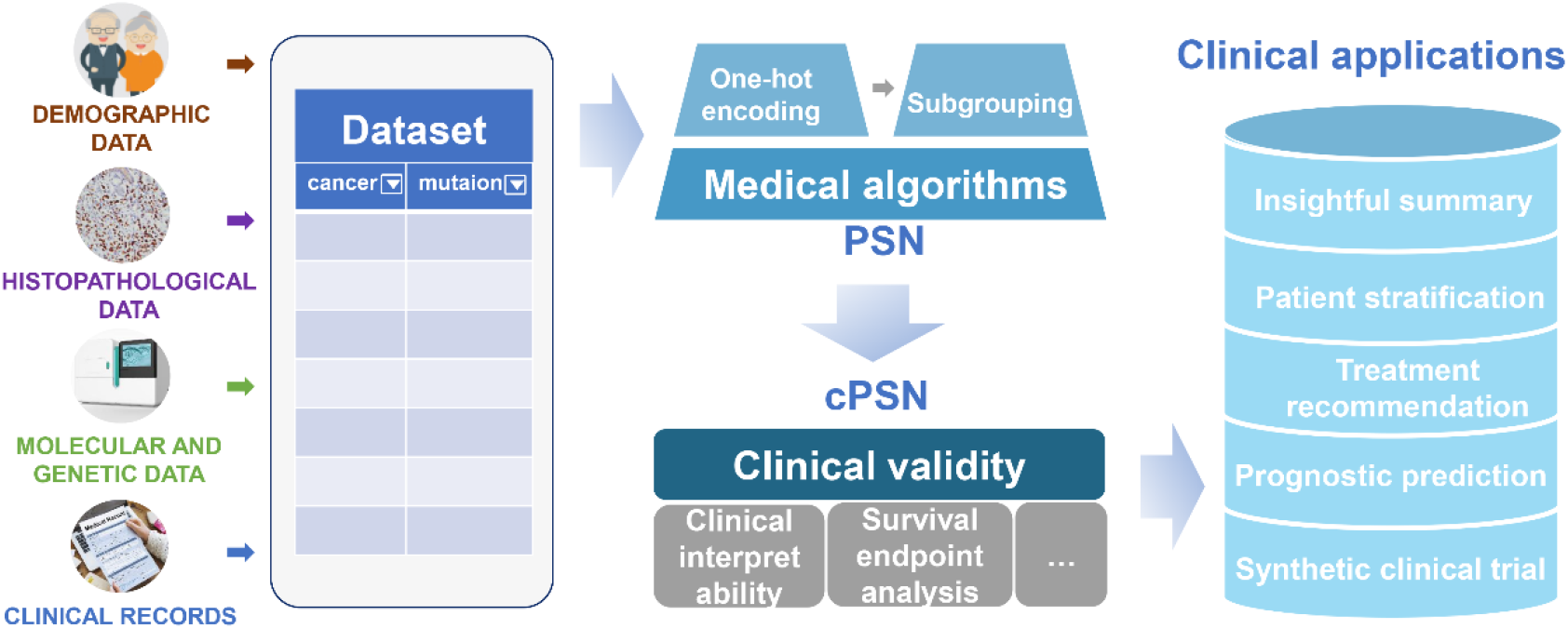
The framework architecture used to construct cancer-specific patient similarity network.

Categorical data representation has advantages in capturing data from clinical records[24]. Numerical data is continuous value that is accurate, but it doesn’t necessarily have to be presented this way. Given continuous value within a certain range could be considered similar clinical significance, and to improve the generalization ability of the model, we transformed continuous values into discrete values by equivalent partitioning. In this case, categorical variables were directly coded in the encoding process, numerical variables and clinical qualitative descriptions were first converted into categorical variables. In order to integrate multimodal medical data, we encoded feature parameters of each patient by engaging one-hot encoding method. A total of 143 one-hot encoding values were identified from 37 variables, as a result of each categorical state of each variable is recorded as a one-hot feature. Subsequently the heterogeneous data of patients were transformed into a feature embedding matrix.

Through feature coding, patients embedding and distance calculation, all patients were orchestrated to form a PSN, which is a *M*-dimensional network where *M* is the sum of observation parameters. PSN reflected the similarity distance between patients (Figure 2). Each pot in the high-dimensional PSN represents a patient. We then conducted cluster analysis. The 1,000 surgical gastric cancers were divided into 2 to 11 clusters via *K*-means algorithm. Learning from elbow method, 5 clusters represent appropriate partition with the best clustering performance. Each cluster represents a similar group, sharing some clinical characteristics (Figure 3), which is the immanent foundation for treatment recommendations for a given patient who is clustered into a specific group.

**Figure 2.**
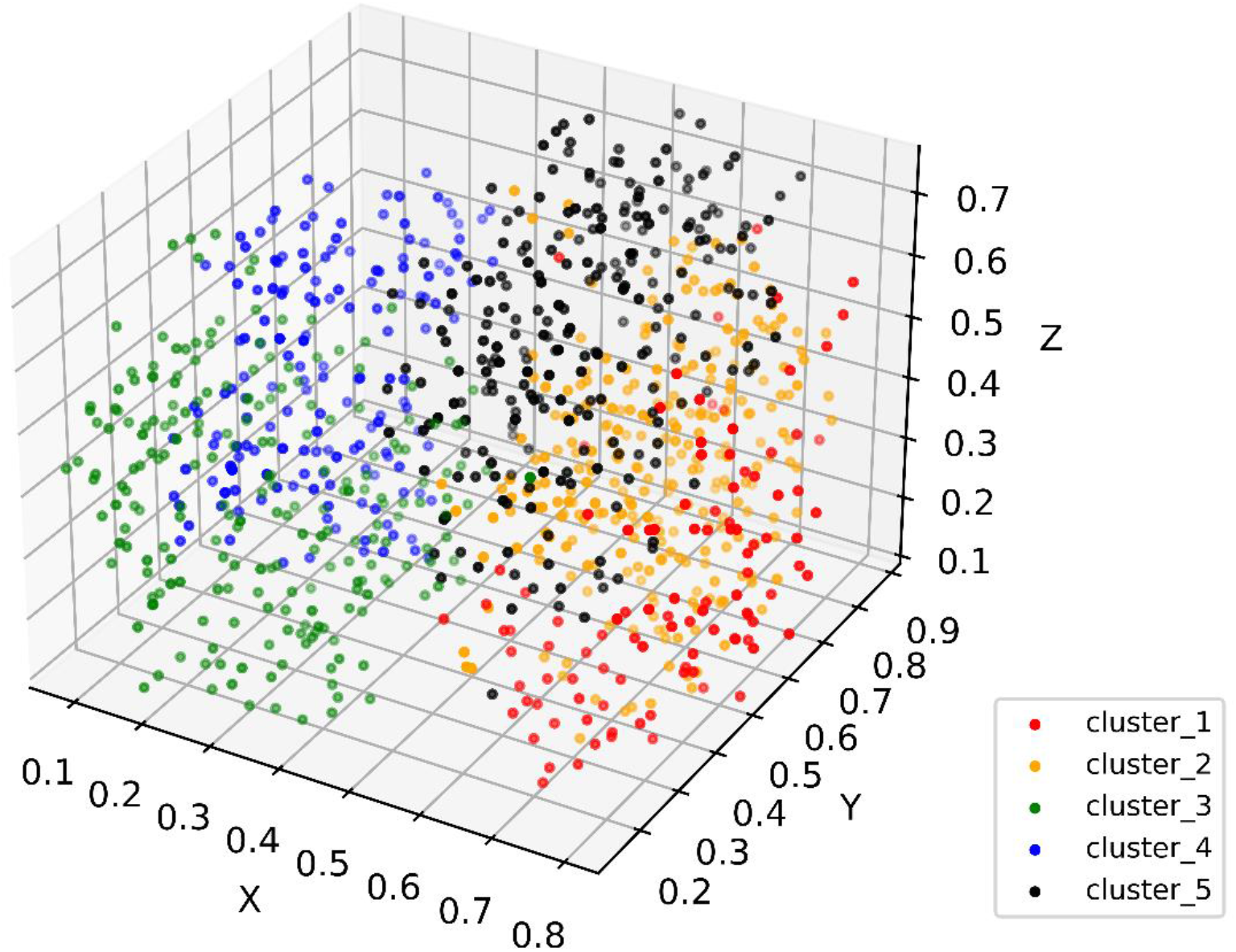
3D *t*-SNE show patient distribution in the constructed cPSN. Colors show different subgroups identified in patient similarity analysis. Values in axes represent relative distance in the dimension.

**Figure 3.**
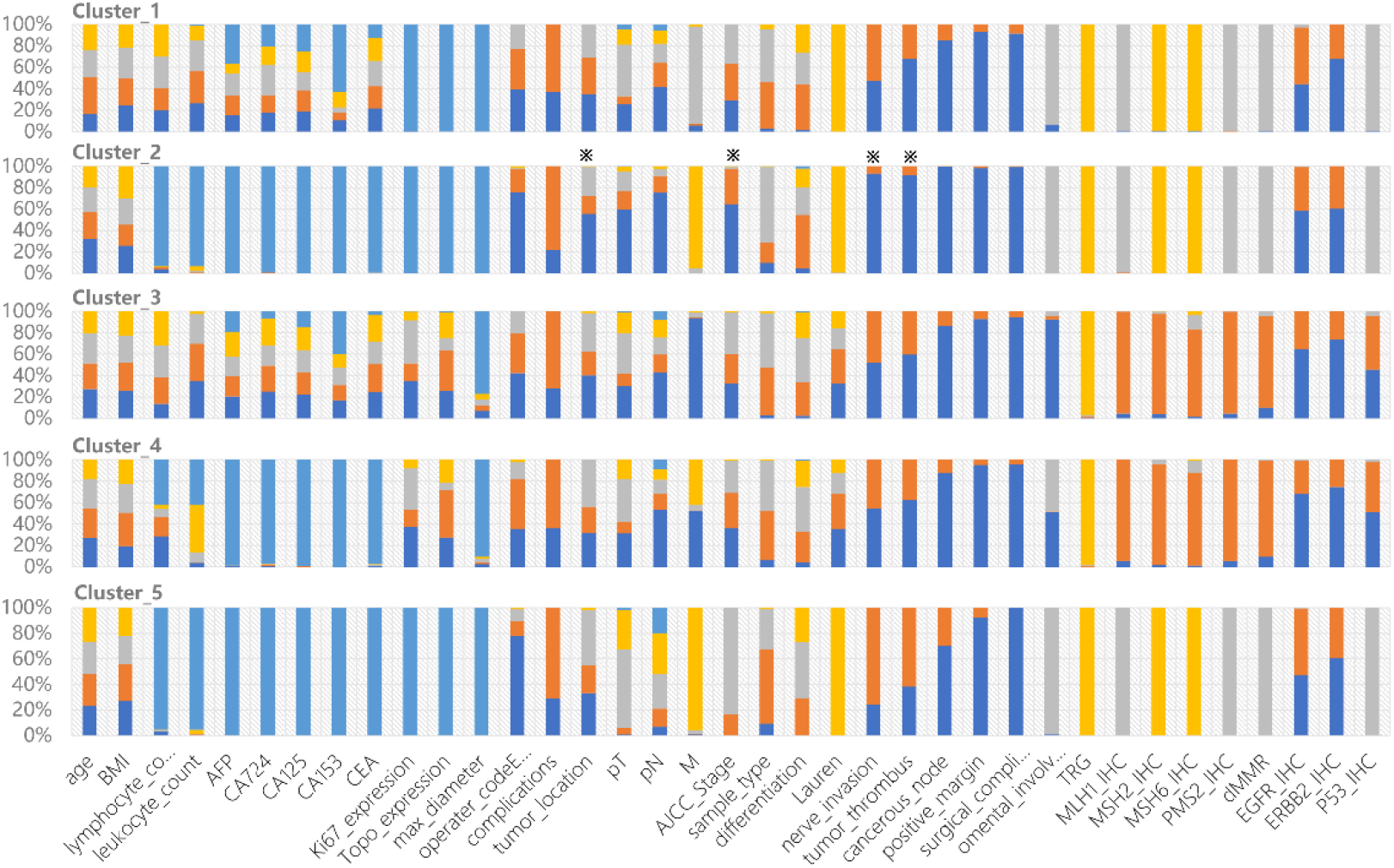
Clinical characteristics of each cluster. The colors show frequency of each categorical state of the variable. All of 37 variables are show in different clusters, respectively. ※ indicate specific features of cluster_2 compared to other clusters.

We performed the correlation analysis of clinically meaningful endpoint to evaluate the clinical validity of the clustering. Overall survival, which served as the gold standard of oncological clinical endpoint[25], was investigated to assess the validity and clinical relevance of the constructed PSN. When the patients in our cohort were divided into five clusters, the OS differences between clusters were statistically significant (log-rank test, *p* <2e-16, Figure 4A). Our strategy achieved excellent performance that was superior to the traditional classifications, such as patient age, cancer differentiation, or tumor stage (Figure 4B,4C,4D).

**Figure 4.**
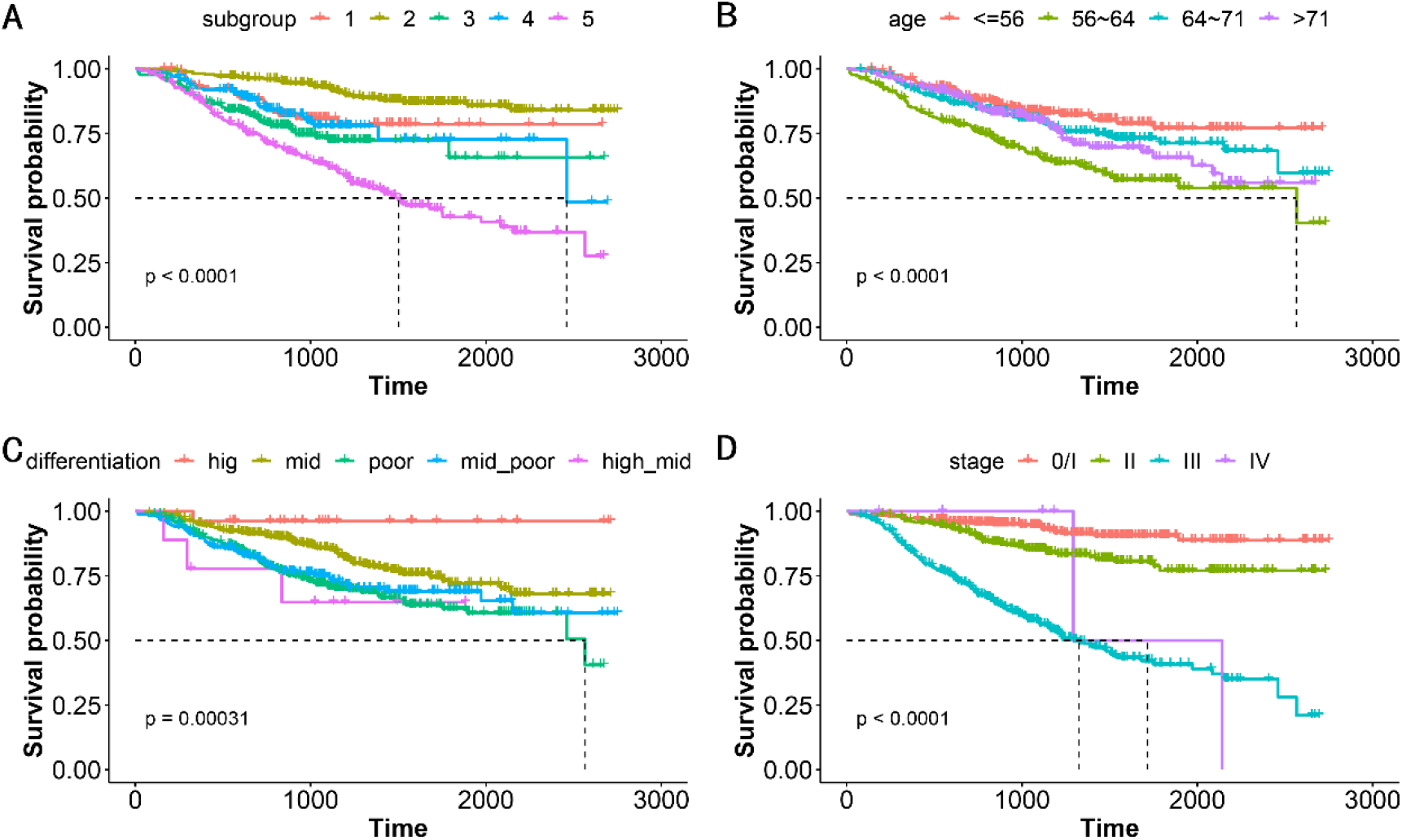
Kaplan–Meier survival analysis for OS by (A) subgroups, (B) patient age, (C) cancer differentiation, and (D) tumor stage. The five subgroups represent the patients are classified 5 clusters based on patient similarity calculation. Patient age is quartile classification. *p*-value shows statistical significance by log-rank analysis.

Cluster_2 had the longest survival rates. Most patients in the cluster_2 are negative nerve invasion, negative tumor thrombus, negative cancerous node, or/and no regional nodal involvement. The patients’ pathological stage is mainly stage I and stage II, although scattered across all stages. All of these clinical indicators support better prognosis. Interestingly, we found that lower distal GCs are more than upper proximal GCs, shedding light on the debates[26, 27] (Figure 3). Cluster_5 had the worst prognosis among all subgroups. The patients’ pathological stage is mainly stage III, and upper tumor location is in the majority. Cluster_1 contain 71.6% of patients of Mx that means distant metastasis cannot be determined. TP53 mutation is dominantly in Cluster_3 and Cluster_4, in accordance with their dMMR characteristic (Figure 3).

## Discussion

The present research provides a similarity calculation method for tumor patients based on one-hot encoding unsupervised clustering. According to their clinical features, a cohort of tumor patients were embedded in a high-dimensional space, then patients are clustered into several groups that share a lot in common. Do these different groups of patients are clinically different? While death is the primary event of interest in cancer patients, based on the OS of cancer patients, the correlation analysis of the clinical endpoint was carried out on clustered patients. The log-rank test assessed statistical significance to examine whether the distribution of OS was distinguishable, which ensures the clinical significance and clinical practical value of the established PSN. For example, the cancer stage is conventionally used to stratify patients[28]. However, patients with different stages were often clustered into the same subset in our model. Furthermore, patients in the same subset are similar in survival prognosis, probably as well as other clinical characteristics and respond to treatment.

Clinical data resources include electronic medical records, imaging examinations, laboratory tests, genetic and cellular analyses. How to integrate patients’ data that are highly heterogeneous is vital in the patient similarity analysis. We adopted the “early integration strategy” that constructs a unified model for all types of data, in contrast to the “late integration strategy” calculates distances for each data type that requires searching for corresponding appropriate model[29]. Note that “early integration strategy” ignore the correlation between parameters. The Mahalanobis distance calculation that weights the multivariate parameter using the covariance matrix may compensate for the shortcoming. Data encoding emerged following integration. We used one-hot encoding strategy as it is concise and robust in clinical data management. It can efficiently code any clinical data, and the data processing ability is outperforming. As the parameters with clinical meaning and data accessibility are limited, dimensionality should never be in mind. Although undesirable, missing data frequently occurs in real-world healthcare scenarios due to the values of the variables are not measured or unavailable for a patient. The usual practice is filling in with estimating values that underrepresent the real state and therefore is not suitable for further analysis. The present study regarded the missing value as an independent one-hot coding type without filling in null values, which may reduce value bias and avoid the classification error caused by filling methods.

Beside multimodal medical data integration and encoding, data labelling lacks standards in the field of tumor patient similarity. Doctor’s annotating only make judgement based on a fraction of information, usually by rule of thumb. These labelling processes are subjective or otherwise uncertain. However, AI algorithms and machine learning usually start with a mount of labelling data. This is a great gap between manual labelling given and accurate labelling demand that is a critical step for training the algorithm. Unsupervised clustering, independent of any labelling data, efficiently classifies patients into subgroups. Then the machine learning can be used to uncover clinical characteristics or data features underlying the subgroups. Essentially, the constructed cPSN should authentically restore the similarity of patients in real-world, linking to prognostic assessment, personal treatment and health management.

While consensus on which machine learning algorithm performs better on specific data types in the context of precision medicine is still lacking[30], the present study performs K-Means unsupervised clustering and evaluates K by statistical algorithm to obtain the optimal K, and the whole process is unsupervised without human intervention. The present study uniformly adopts one-hot encoding for multi-modal, highly heterogeneous clinical data and is flexibly compatible with clinical data evolvement and changes in observation status caused by different medical institutions, doctors, and medical development stages. The data processing method provides an extensibility mechanism for adding more parameters. In future, when synthetic data is expected to replace real data in medical big data analytics[31], machine learning algorithms can be used to mine data, which is a subsequent mission of patient similarity analysis.

Altogether, we developed an easy-to-perform, clinically interpretable, generalizable and universal method to conduct cancer patient similarity analysis. For a target patient to be evaluated, a group of patients most similar to the target index patient was obtained in cPSN using the K-nearest neighbor algorithm based on distance calculation, and the range and fineness of the similar patients to the index patient were selected by adjusting the K value. This is the way in where population-based clinical information, searching similar patient cases for proposing treatment and management strategies, which would promote the development of big-data-based precision medicine.

This study may have several limitations so that the results must be interpreted with caution. Although one-hot encoding is a robust method that could dispose of missing values, too many null values in the dataset affect the accuracy of the results. It is better to apply as complete records as possible in future studies. Besides, the data we used were baseline data that depicted the patients’ features before surgery, without considering treatment information. That may make sense because the causal connection exists between baseline data and treatment programs. Our model ignored the correlations among the parameters of selected features. We keep in mind that parameter redundancy and non-weight matrices are unconformity of the cPSN. These require solutions, especially under the framework of unsupervised learning.

## Conclusions

Through integrating heterogeneous clinical data (e.g. histopathological data, molecular and genetic data, laboratory data, imaging data), we constructed a clinical patient similarity network (cPSN). The constructed cPSN model is scalable, generalizable, and performs well for various data types. Moreover, our cPSN is associated with clinical implications, which would give researchers insights towards clinical issues, and could help clinicians make treatment decision, guide clinical management and predict clinical outcome.

## Data Availability

All data produced in the present study are available upon reasonable request to the authors

## Data Availability

The data used to support the findings of this study are available from the corresponding author upon reasonable request.

## Ethics Approval and Consent to Participate

This study was approved by the Ethics Committee of Changhai Hospital. Informed consent was obtained from all patients for the study.

## Conflict of Interest Disclosures

The authors declare no conflicts of interest.

## Funding

This work was supported by the National Natural Science Foundation of China (#91846302).

## Notes

### Competing Interest Statement

The authors have declared no competing interest.

### Author Declarations

This study was approved by the Ethics Committee of Changhai Hospitall, Naval Military Medical University.

